# Tobacco Use and Associations with Alcohol Consumption and Socio-Demographic Factors Among Cambodian Men Aged 15–49 Years: Evidence from the Cambodia Demographic and Health Survey 2021–2022

**DOI:** 10.1101/2025.06.04.25328721

**Authors:** Samnang Um, Leang Supheap, Pall Chamroen, Chantrea Sieng, Sovandara Heng, Grace Marie Ku

## Abstract

Tobacco use remains a leading preventable cause of illness and death globally, contributing to significant public health, environmental, and socio-economic challenges. This study aimed to investigate associations of tobacco use with alcohol consumption and socio-demographic factors among Cambodian men aged 15–49 years. This study used men’s data from the Cambodia Demographic and Health Survey 2021-2022. A total of 8,825 men aged 15–49 years were included in the final analysis. The outcome variable was ‘tobacco use’, which was further categorized into smoked tobacco and smokeless tobacco use. Multiple logistic regression was performed to identify determinants of tobacco use, smoking tobacco, and smokeless tobacco use. The overall prevalence of any tobacco use, smoked tobacco, and smokeless tobacco use was 21.5% (95% CI: 20.2–22.9), 20.1% (95% CI: 18.9–21.4), and 0.93% (95% CI: 0.7–1.2), respectively. The highest rates of any tobacco use were observed in Stung Treng (40.7%), followed by Mondulkiri and Ratanak Kiri, each at 39%. Similarly, the prevalence of smoked tobacco is highest in Mondul Kiri (36.5%), Ratanak Kiri (34.5%), Stung Treng (32.5%), and Kratie (30.4%). Meanwhile, smokeless tobacco use is relatively low across most provinces, with the highest rates in Mondulkiri (19.1%), Stung Treng, and Pailin (each at 6%). Several independently significant predictors of any tobacco use, smoked tobacco use, and smokeless tobacco use were higher in young adult men aged 19–24 years, no or limited education, agricultural/domestic and unskilled or skilled manual jobs, poor wealth, and alcohol consumption. Public health promotion programs need to strengthen tobacco control, particularly among the younger age groups, and can take inspiration from successful tobacco control initiatives implemented in other countries. Social determinants of health, which themselves influence health outcomes, should be addressed. Policies integrating actions to control both tobacco and alcohol can be considered.

## Introduction

Twenty years after the World Health Assembly’s adoption of the Framework Convention on Tobacco Control (FCTC), tobacco use remains a major risk factor causing preventable illness and death worldwide, contributing to public health crises, environmental degradation, and significant economic losses [1–3]. In 2023, the World Health Organization (WHO) reported that tobacco use is responsible for over 8 million deaths annually, with 1.3 million attributed to secondhand smoke exposure [2,4]. Tobacco-related diseases, including cancers (lung, head, neck, esophageal, and bladder), cardiovascular conditions, diabetes, and chronic respiratory diseases, disproportionately affect low- and middle-income countries (LMICs), where approximately 80% of the world’s 1.3 billion tobacco users reside [2,4,5]. Globally, 22.3% of the population use tobacco, with higher prevalence among men (36.7%) than women (7.8%) [2].

Despite increasing awareness of the risks and tobacco control in place [6], tobacco use remains widespread in Southeast Asia, where socio-economic and cultural factors, combined with the accessibility of tobacco products, sustain high consumption rates [7,8]. For instance, smoking prevalence among adults aged 15 and older in Thailand in 2022 was 19%, with 37.7% of men and 1.8% of women identified as smokers [9]. Youth aged 10-14 reported a smoking prevalence of 7%, with boys at 11.3% and girls at 3.3% [9]. The leading causes of death in 2021 in Thailand were stroke, ischemic heart disease (IHD), lung cancer, and chronic obstructive pulmonary disease (COPD) ranking first, second, sixth, and tenth, respectively. Tobacco use significantly contributes to these diseases, accounting for 10.6% of all deaths, with a higher impact on men (15.6%) than women (4.3%) [9]. Tobacco was listed as the fourth highest risk factor driving disability and deaths in the Philippines in 2021 [10]. Cambodia, like other countries in the region, faces a significant public health challenge due to widespread tobacco use, undermining efforts to achieve national and global health objectives, including the Sustainable Development Goals (SDGs) [11–13].

In Cambodia, tobacco-related diseases contribute to over 15,000 deaths annually, and the economic costs due to healthcare spending and lost productivity exceed $663 million [14]. The Royal Government of Cambodia ratified the WHO FCTC on November 15, 2005 [15]. Cambodia’s Tobacco Control Law, enacted in 2015, laid the foundation for tobacco control efforts, including establishing 135 cessation and counseling centers nationwide. A 2016 sub-decree, in collaboration with the Ministry of Tourism, banned smoking in public spaces, leading to the certification of 511 smoke-free tourism establishments by 2022. However, despite these efforts, a 2023 WHO survey revealed that 35.5% of men and 2.4% of women aged 18-69 continue to use tobacco [16]. Furthermore, 16% of adults report exposure to secondhand smoke, especially in public spaces like restaurants (51%) and public transport (62%) [16]. The rise in electronic cigarette use among the youth adds a new challenge, with 82% of vape users aged 15-35, and over 80% of these users purchasing products online [17]. The increasing use of electronic smoking devices, particularly among the youth, presents a new public health concern. In response, the Cambodian Ministry of Health is currently working on a sub-decree to ban the sale and use of electronic cigarettes and shisha.

The 2021 Cambodia National Adult Tobacco Survey (NATS) provides a detailed picture of tobacco use in Cambodia, revealing that 2.17 million adults use tobacco, including 1.63 million smokers. Among men, 28.62% use tobacco, with 25.35% identified as smokers, while 8.88% of women use tobacco, and 2.05% smoke. Tobacco use is higher in rural areas (14.34%) than in urban areas (10.73%) [17]. The economic burden is evident, with tobacco users spending $235 million annually on tobacco products and rural users spending more than their urban counterparts [17]. These findings underscore the urgent need for targeted tobacco control strategies to address Cambodia’s health and economic burdens.

According to the STEPS survey country report in 2023 in Cambodia, among men aged 18-69, 35.8% reported using tobacco, including both smoking and smokeless forms. Tobacco use was more prevalent among older men aged 45-69, with a rate of 42.2%. Among those who self-reported their alcohol consumption, 71.2% of men aged 18-69 had consumed alcohol in the past 30 days. Studies demonstrated a direct association between tobacco use and alcohol consumption in both adults and youth [18–21], and tobacco-alcohol interactions increase risks for non-communicable diseases such as hypertension [20]. However, alcohol control policies are not as advanced as tobacco control not only in LMICs but also in HICs [22]. A more nuanced analysis looking into socio-demographic and behavioral factors, including alcohol use, associated with tobacco use has not yet been done and will provide more evidence for more precise targeting of tobacco control and the possibility of integrating alcohol control strategies in Cambodia. The analysis likewise provides evidence on the significance of social determinants not only on health and well-being but also on health risks.

## Study Objective

The study aimed to examine the socio-demographic and behavioral factors, including alcohol consumption associated with tobacco use among Cambodian men aged 15–49 years, using data from the Cambodia Demographic and Health Survey 2021–2022. The findings will contribute to the development of targeted tobacco control as well as alcohol control strategies and public health policies aimed at reducing tobacco-related and alcohol-related health disparities and mitigating the broader public health impact of tobacco and alcohol use in Cambodia in particular, and in addressing social determinants of health in general.

## Methods

### Ethics statement

This study analyzed men’s data extracted from the most recent Cambodia Demographic and Health Survey (CDHS) 2021–2022, which is publicly accessible upon request through the DHS Program website (https://dhsprogram.com/data/availabledatasets.cfm). All personal identifiers were removed to ensure participant confidentiality. The dataset contains no individual identifiers that could compromise the privacy of the participants, and the data were used solely for analysis purposes. Written informed consent was obtained from the parent/guardian of each participant under 18. The protocol for the CDHS 2022 was approved by the Cambodia National Ethics Committee for Health Research on May 10, 2021 (**Ref: 083 NECHR**) and by the Institutional Review Board (IRB) of ICF International in Rockville, Maryland, USA.

### Data sources and procedures

The Cambodia Demographic and Health Survey (CDHS) 2021–2022 was a nationally representative household survey conducted between September 2021 and February 2022, with a temporary pause due to the COVID-19 pandemic. The primary goal of the survey was to provide updated and reliable estimates on a wide range of demographic and health indicators, including fertility, family planning, breastfeeding, nutrition, maternal and child health, adult and childhood mortality, women’s empowerment, domestic violence, and awareness of HIV/AIDS and other sexually transmitted infections (STIs). Additionally, the survey collected data on health-related behaviors such as smoking, alcohol use, and consumption of unhealthy foods [23].

The survey employed a two-stage, stratified cluster sampling design, using the Cambodia General Population Census of 2019 as its sampling frame [24]. In the first stage, 709 enumeration areas (EAs) were selected, comprising 241 urban and 468 rural areas. In the second stage, a systematic sample of 25 to 30 households was drawn from each EA, totaling 21,270 households. The household response rate was exceptionally high at 99% [23]. Of the above, 8,825 men aged 15–49 were interviewed from half of the selected households, with a response rate of 97%. Trained interviewers conducted face-to-face interviews using standardized questionnaires to ensure the consistency and accuracy of the data collected [23]. For additional details on the survey methodology, sampling procedures, and data collection process, readers can refer to the final CDHS report [23].

### Study participants

The data of all male respondents in the CDHS 21-22 survey were included in the analysis.

### Measurements of variables

#### Outcome variables

The study focused on **tobacco use** among men aged 15–49 years, categorized into three groups:

1. **Any tobacco use** (1 = user, 0 = non-user),
2. **Smoked tobacco use** (1 = smoker, 0 = non-smoked tobacco use),
3. **Smokeless tobacco use** (1 = smokeless tobacco, 0 = non-smokeless tobacco use),

A participant was considered a smoker if he reported using any form of smoked tobacco, including manufactured cigarettes, hand-rolled cigarettes, kretek cigarettes (clove-flavored), tobacco pipes, cigars, cheroots, cigarillos, water pipes, or other smoked tobacco products at the time of the survey. Those who reported using snuff (by mouth or nose), chewing tobacco, or betel quid with tobacco were categorized as users of smokeless tobacco. Participants who reported using either smoked or smokeless tobacco were classified under any tobacco use [23].

#### Independent Variables

Independent variables consisted of sociodemographic factors, namely: age groups (15–18, 19–24, 25–29, 30–39, and 40–49 years); marital status (not married, married or living together and divorced or widowed or separated); parity (no children, one-two children, three-four children, and five or more children); educational level (no education, incomplete primary, completed primary, incomplete secondary, completed secondary, and higher); occupation (not working, agricultural/domestic, professional/managerial, clerical/sales/services, and skill/unskilled manual labor); health insurance coverage (coded as no and yes), including public and private insurance; and the household wealth quintile, which was calculated using principal component analysis (PCA) with variables for household assets and dwelling characteristics, and then divided into five groups: poorest, poorer, middle, richer, and richest [23,25,26].

Other independent variables include household information: relationship to the head of the household (coded as Head of Household, Spouse/Partner/Parent, Child (Son/Son-in-law/Grandson), Sibling (Brother), Other Relative, and Non-relative/Care); household size (coded as small (≤ 3), medium (4–6), and large (≥ 7); years of cohabitation (coded as never married, 0–4 years, 5–9 years, 10–14 years, 15–19 years, 20–24 years, 25–34 years).

Variables related to behaviour include: alcohol consumption was classified as non-drinker and drinker and frequency per month (non-drinker, drinks only on 1–5 days per month, on 6–10 days, on 11–24 days, and daily/almost daily for those who drink 25 days or more per month). Reading a magazine or newspaper, listening to the radio, and watching television were coded as no and yes. Mobile ownership was categorized as (no mobile telephone, non-smartphone, and smartphone). The frequency of internet use in the last month was categorized as (never, less than once a week/at least once a week, and daily/almost daily). And self-reported health status was categorized as good, moderate, or poor.

Further data collected comprised the participant’s place of residence (rural vs urban), geographical location coded as one of five geographical regions (the capital Phnom Penh, Plains, Tonle Sap, Coastal/sea, and Mountainous), and religion (Buddhist, Muslim/Christian) as defined in Cambodia’s General Population Census 2019 [24].

### Statistical analysis

The data was analyzed using STATA V18 (Stata Corp, Texas, 2023) [27]. The standard DHS sampling weight and complex survey design were accounted for using the survey package in the descriptive and logistic regression analyses. Descriptive statistics were used to estimate key socio-demographic and behavioral factors, with results presented as weighted frequencies and percentages. The prevalence of tobacco use and patterns among men was also estimated by province separately, and categorized into the three groups (any tobacco use, smoked tobacco use, and smokeless tobacco use). These were visualized using ArcGIS software version 10.8 [28]. The administrative boundaries of Cambodia’s shapefile were obtained from the United Nations Coordination of Humanitarian Affairs (https://data.humdata.org/dataset/cod,ab,khm) [29]. A bivariate analysis using chi-square tests was conducted to explore associations between socio-demographic and behavioral factors with tobacco use. Variables that showed a significant association (p-value ≤ 0.05) in the bivariate Chi-saure analysis were then included in the final multiple logistic regression analysis [21]. The multiple logistic regression model was used to identify independent factors associated with tobacco use, adjusting for potential confounders and presenting the results as adjusted odds ratios (AOR) with 95% confidence intervals (CI).

### Model Diagnostics for Multiple Logistic Regression

To check for multicollinearity, we calculated the Variance Inflation Factor (VIF) for each independent variable included in the logistic regression models. These variables were: years of cohabitation, age, marital status, wealth index, education level, mobile phone ownership, internet use, newspaper reading, radio listening, television watching, place of residence, geographic region, alcohol consumption, and occupation. We considered VIF values above 5 as a sign of possible multicollinearity and values above 10 as indicating a more serious issue. All variables had VIFs below 5, suggesting that multicollinearity did not pose a problem for the analysis (**S1 Table)**. Model fit was evaluated using the Hosmer-Lemeshow test, and the pseudo R-squared values (Cox & Snell, Nagelkerke) were examined to assess explanatory power. Goodness-of-fit tests were performed for all logistic regression models (**S2 Table**). To assess discrimination, Receiver Operating Characteristic (ROC) curves were generated, and the area under the curve (AUC) was reported for each model . Additionally, interaction terms to predicted probabilities of any tobacco use by interaction effects: Age and Education; Age and Alcohol Consumption; Education and Occupation; Mobile Phone Ownership and Internet Use.

## Results

### Socio-demographic characteristics and behaviors

The study included 8,825 men with a mean age of 30.5 years (SD 9.6). The largest age group is 30–39 years (32.3%), followed by 40–49 years (21.6%). Most participants are married or living with a partner (63.5%), while 33.1% are non-married. 37.5% had incomplete secondary education, 29.5% had incomplete primary education, and 7.7% had no formal education. Most participants were employed in skilled or unskilled manual labor (37.6%) or agricultural/domestic roles (32.6%). Over 26.0% belonged to the poorest household, whereas 16.5% were in the richest. Health insurance coverage was low, with 87.4% reporting no coverage. Most participants resided in medium-sized households (4–6 members, 63.3%), while 20.7% lived in small households. Urban residence was reported by 64.1% of participants, with 30.2% living in the Tonle Sap region. Approximately 74.0% of participants did not read newspapers or magazines, 79.8% did not listen to the radio, and 57.4% did not watch television. Smartphone ownership was 82.2%, and 63.8% reported daily internet use. Most participants rated their health as good (68.4%), with 29.6% reporting moderate health and 2.0% poor health. Alcohol consumption was prevalent, with 71.0% reporting alcohol use in the past month. Of these, 44.0% consumed alcohol on 1–5 days in a month, 13.8% on 6–10 days, and 6.0% consumed alcohol daily or almost daily (see **Table 2**).

### Prevalence and pattern of tobacco use

The overall prevalence of tobacco use in any form was 21.5% (95% CI: 20.2–22.9). Smoked tobacco products use was 20.1% (95% CI: 18.9–21.4), and smokeless tobacco use was 0.93% (95% CI: 0.7–1.2), respectively. Among users of smoked tobacco products, most (87.3%) consume fewer than five cigarettes per day, while smaller proportions smoke 5–9 cigarettes (2.6%), 10–14 cigarettes (3.9%), 15–24 cigarettes (5.3%), or more than 25 cigarettes (0.9%). Commercially manufactured cigarettes are the most frequently used product (17.6%; 95% CI: 16.3–18.9), followed by hand-rolled cigarettes (3.2%; 95% CI: 2.8–3.8) and kreteks (1.9%; 95% CI: 1.6–2.3), and among the least used are pipes (1.7%; 95% CI: 1.4–2.1), cigars or cheroots (1.7%; 95% CI: 1.4–2.1), and water pipe sessions (1.7%; 95% CI: 1.4–2.1). Regarding types of smokeless tobacco, snuff by mouth (0.5%), snuff by the nose (0.3%), chewing tobacco (0.2%), betel quid with tobacco (0.2%), and other types of smokeless tobacco (0.2%) are reported (see **Table 1**).

**Table 1.** Patterns of difference tobacco use among men in Cambodia during the survey period 2021–2022 (N=8,825)

| **Outcomes** | **Freq.** | **Percent** | **95% CI** |
| --- | --- | --- | --- |
| **Any tobacco use** |  |  |  |
| Non-user | 6,924 | 78.5 | 77.1–79.8 |
| User | 1,901 | 21.5 | 20.2–22.9 |
| **Smoked tobacco** |  |  |  |
| Non-smoker | 7,050 | 79.9 | 78.6–81.1 |
| Smoker | 1,775 | 20.1 | 18.9–21.4 |
| **Smokeless tobacco** |  |  |  |
| Non-smokeless | 8,743 | 99.1 | 98.8–99.3 |
| Smokeless | 82 | 0.93 | 0.7–1.2 |
| **Number of tobacco smoked per day (n=1,775)** |  |  |  |
| < 5 | 7706 | 87.3 |  |
| 5–9 | 230 | 2.6 |  |
| 10–14 | 341 | 3.9 |  |
| 15–24 | 468 | 5.3 |  |
| ≥ 25 | 80 | 0.9 |  |
| **Types of tobacco products used** |  |  |  |
| Manufactured cigarettes | 1550 | 17.6 | 16.3–18.9 |
| Hand rolled-cigarettes | 285 | 3.2 | 2.8–3.8 |
| Kreteks-cigarettes | 168 | 1.9 | 1.6–2.3 |
| Tobacco pipes | 153 | 1.73 | 1.4–2.1 |
| Cigars, cheroots, or cigarillos | 150 | 1.7 | 1.4–2.1 |
| Water pipes | 151 | 1.7 | 1.4–2.1 |
| Others | 156 | 1.8 | 1.4–2.2 |
| **Types of smokeless products used** |  |  |  |
| Snuff, by mouth | 44 | 0.5 |  |
| Snuff, by nose | 27 | 0.3 |  |
| Chewing tobacco | 15 | 0.2 |  |
| Betel quid with tobacco | 15 | 0.2 |  |
| Other type of smokeless tobacco | 15 | 0.2 |  |

**Noted:** Survey weights were applied to obtain weighted percentages.

### The geographical variation distribution of any tobacco use, smoking tobacco, and smokeless tobacco

**Fig 1** presents the prevalence of any tobacco use among men aged 15-49 years across Cambodia’s provinces. The prevalence of any tobacco use is highest in Stung Treng (40.7%), Mondul Kiri (39.2%), and Ratanak Kiri (39.1%), and Preah Vihear (35.2%) (see **Map A**). Similar, the prevalence of smoked tobacco use is highest in Mondul Kiri (36.5%), Ratanak Kiri (34.5%), Stung Treng (32.5%), and Kratie (30.4%) (see **Map B**). Smokeless tobacco use is relatively low across most provinces, with the highest prevalence reported in Mondul Kiri (19.1%) and Stung Treng, Pailin (each 6%) (see **Map C**).

**Fig 1.**
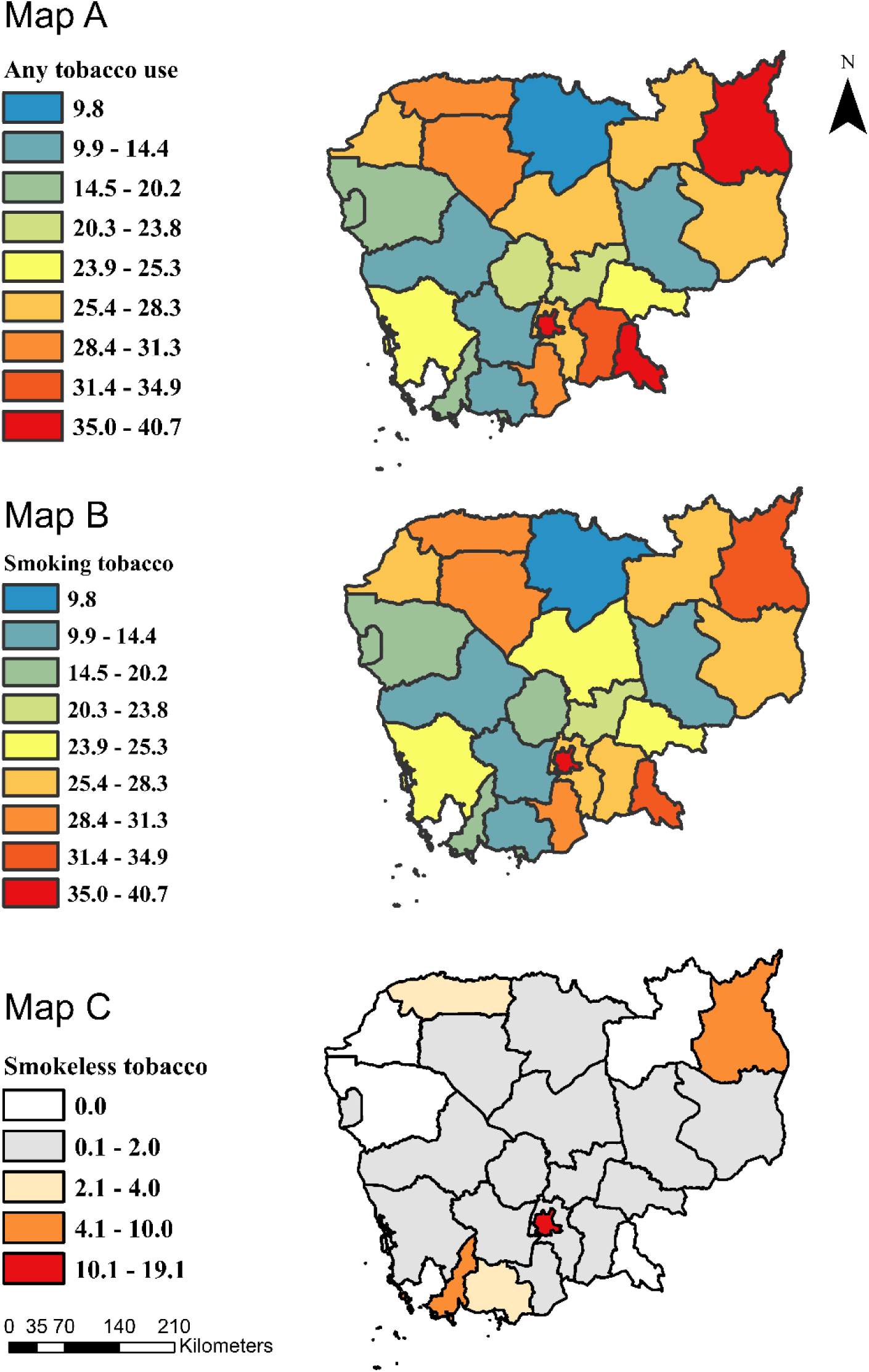
Regional Variation in Tobacco Use: **Map A**: Prevalence of any form of tobacco; **Map B**: Prevalence of smoked tobacco; **Map C**: Prevalence of Smokeless. A shapefile for Cambodian administrative boundaries was obtained from the United Nations for Coordination of Humanitarian Affairs at (https://data.humdata.org/dataset/cod-ab-khm) [29].

### Chi-Square Associated with Any, Smoked, and Smokeless Tobacco Use among Men Aged 15–49 Years in Cambodia

The chi-square analysis identified significant associations between sociodemographic factors and the three categories of tobacco use among the study population.

Any tobacco use was highest among older men aged 40–49 years at 35.7%, compared to the youngest age group (15–18 years) at 5.8% (p < 0.001). Widowed, divorced, or separated men have the highest prevalence (32.2%) (p < 0.001). Men with no formal education had the highest smoking rate (50.5%), while those with higher education had the lowest (1.8%) (p < 0.001). Agricultural workers (30.8%) and manual laborers (22.7%) reported higher smoking rates compared to professional or managerial workers (6.4%) (p < 0.001). Any tobacco use was more common among men from the poorest households (36.9%) (p < 0.001). Any tobacco use was higher among alcohol drinkers (24.9%) compared to non-drinkers (12%) (p < 0.001). Regional differences were observed, with the highest prevalence in the coastal region (27.2%) and the lowest in Phnom Penh (9.8%) (p < 0.001). Additionally, any tobacco use was inversely associated with mobile phone ownership and internet use, particularly among those who owned smartphones and used the internet daily (p < 0.001) (see **Table 2**).

**Table 2.**
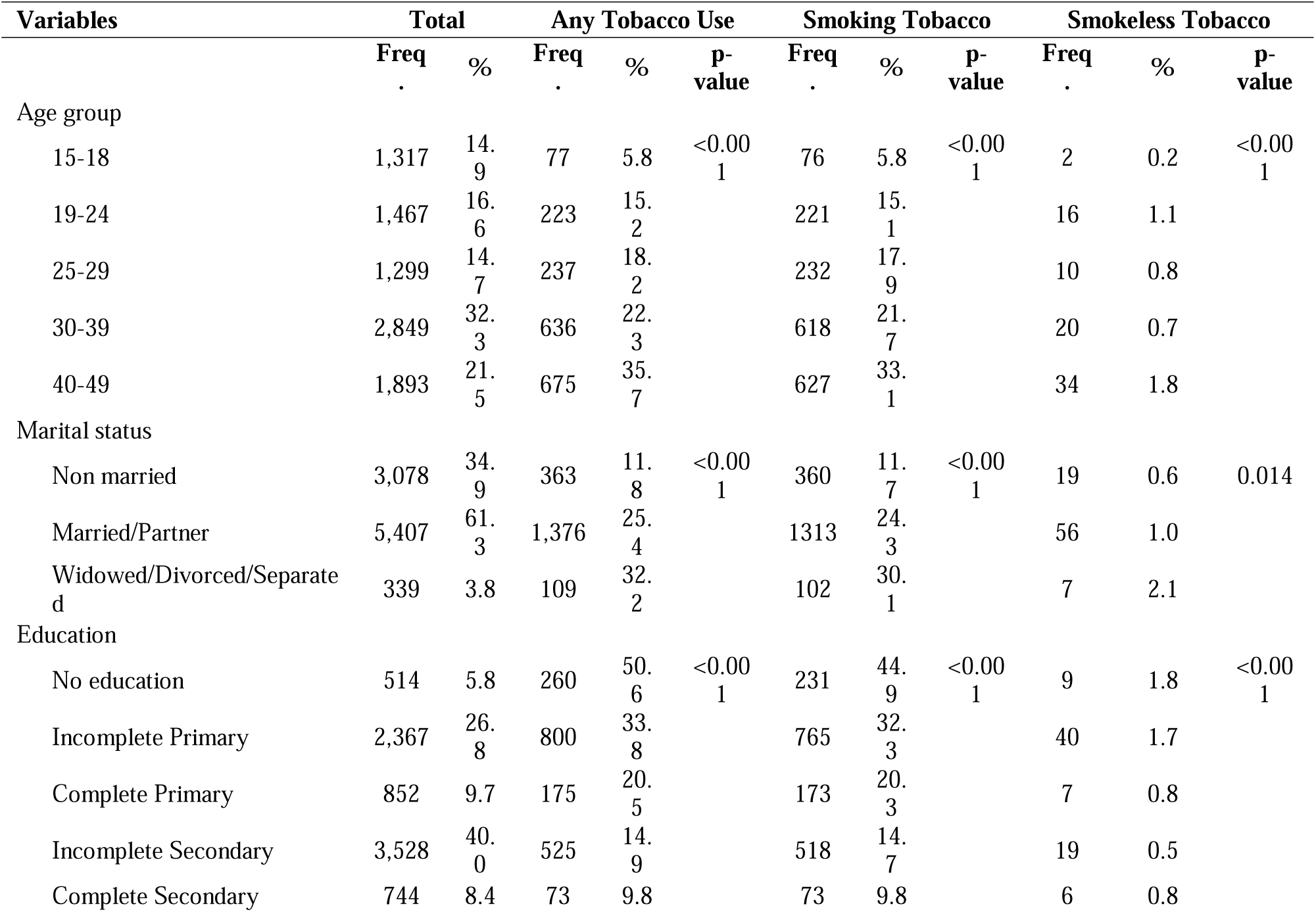
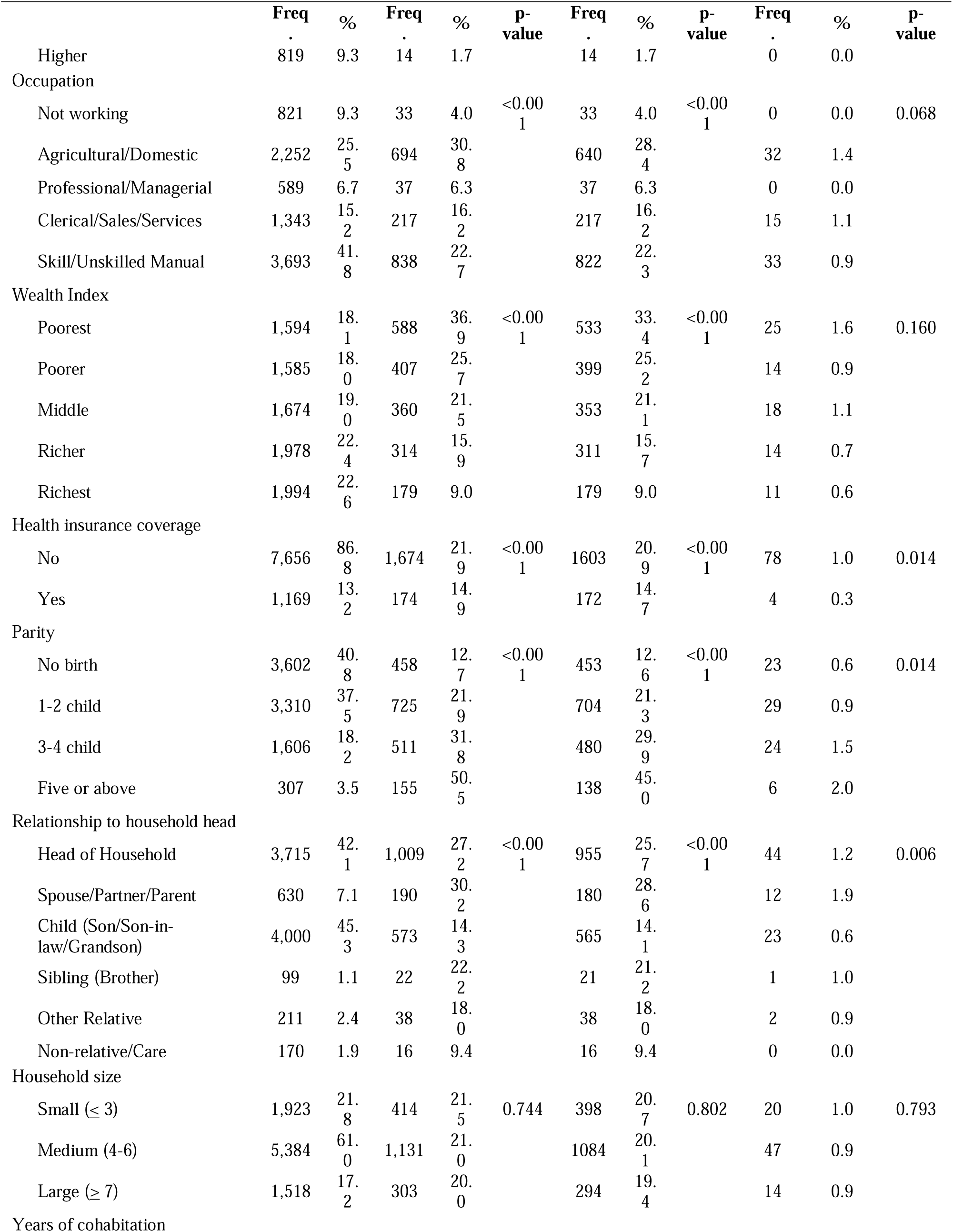
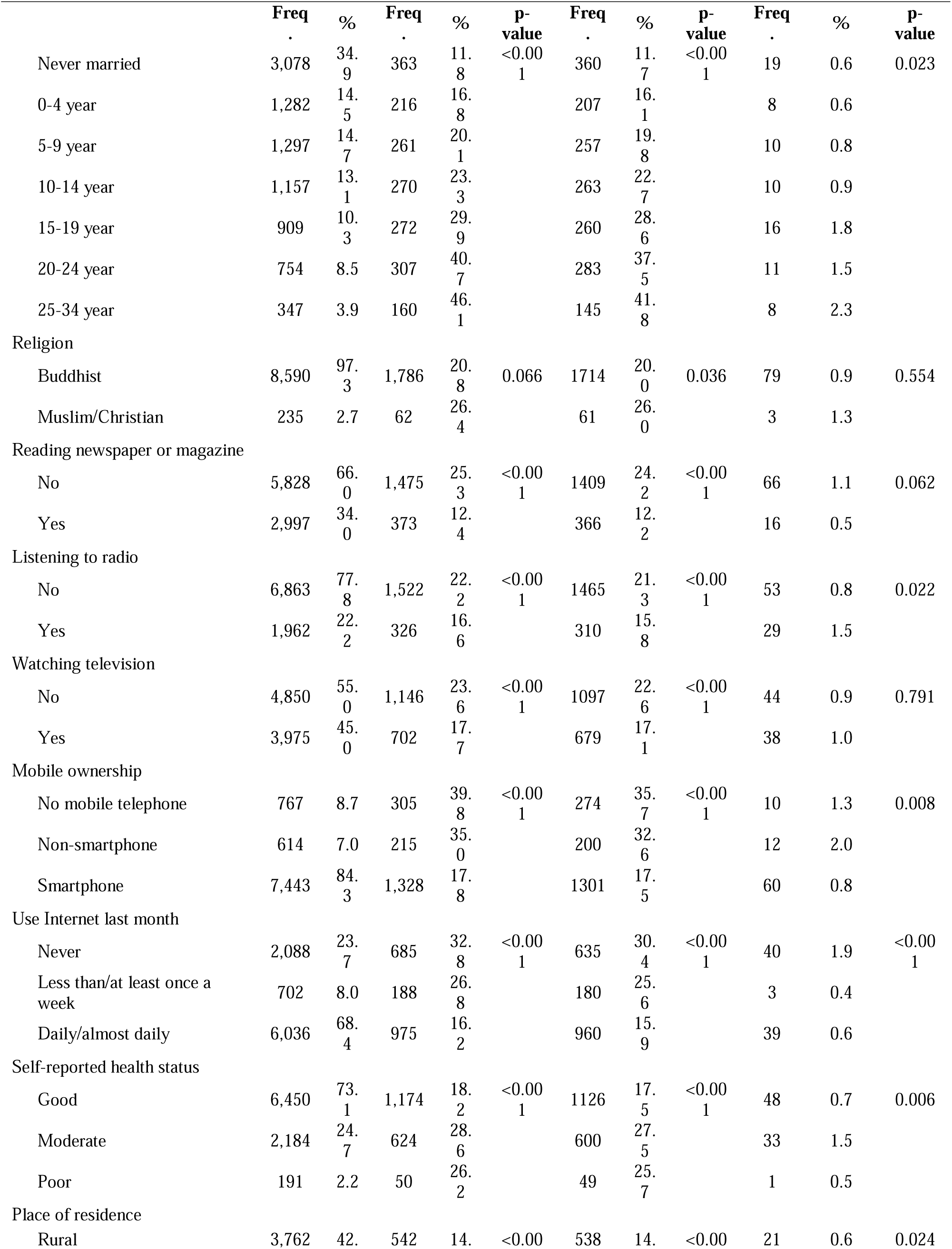
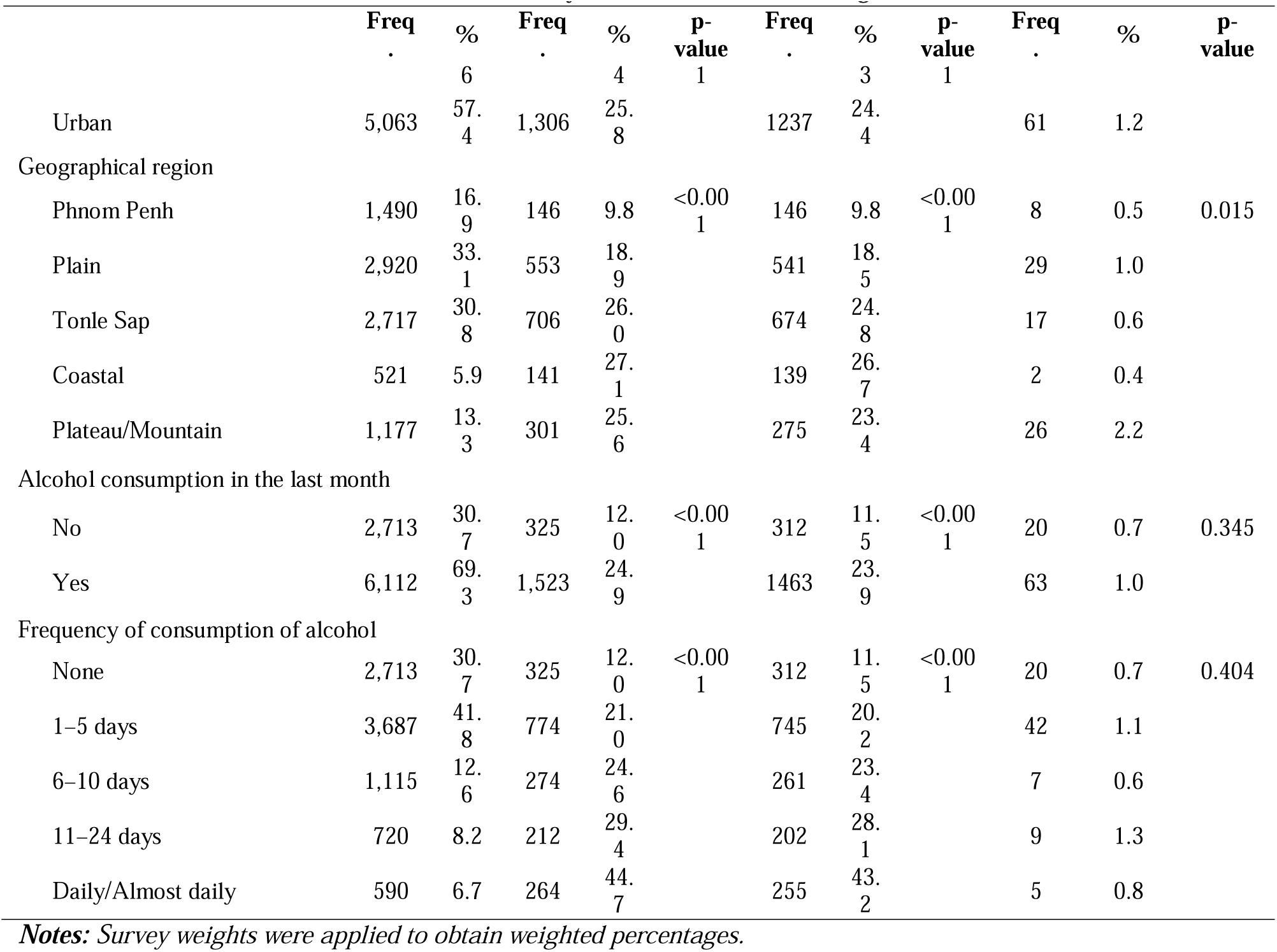
Descriptive statistics and factors associated with any tobacco use, smoking, and smokeless tobacco in chi-square analysis by socio-demographic characteristics and behaviors of men aged 15–49 years (N=8,825)

Similar to the patterns of any tobacco use, smoked tobacco use was most prevalent in older men aged 40–49 (33.1%) compared to those aged 15–18 (5.8%, p < 0.001). Widowed, divorced, or separated men have the highest prevalence (30.1%, p < 0.001). Men with no formal education had the highest smoking rate (44.9%), while those with higher education had the lowest (1.7%, p < 0.001). Smoking tobacco is more common among men from the poorest households (33.4%, p < 0.001). The rate of tobacco smoking was higher among those who drink alcohol (23.9%) compared to non-drinkers (11.5%) (p < 0.001). The same regional differences were observed, with the highest prevalence in the coastal region (26.7%) and the lowest in Phnom Penh (9.8%) (p < 0.001). Additionally, tobacco use was inversely associated with mobile phone ownership and internet use, particularly among those who owned smartphones and used the internet daily (p < 0.001) (see **Table 2**).

Several socio-demographic factors influenced smokeless tobacco use among Cambodian men. Older men, particularly those aged 40-49 years (2.4%), had the highest prevalence, compared to 0.2% among younger men (p < 0.001). Widowed, divorced, or separated men had the highest rates (2.1%), while never-married men had the lowest (0.1%) (p = 0.014). Among those who reported to be married or have a partner, longer cohabitation was associated with higher use, particularly among those in relationships for 25+ years (p < 0.001). Men with no education had the highest use; use decreased with higher education levels (p < 0.001). Agricultural workers had the highest prevalence (1.4%) (p < 0.001). Uninsured men were more likely to use (p = 0.014), as were household heads (p = 0.006). Regional differences and access to technology also played a role, with geographical variations (p < 0.001) and lower use among men with technology access (p < 0.001) (see **Table 2**).

### Adjusted Odds Ratios Evaluating Factors Associated with Any Tobacco Use

We performed multiple logistic regression to evaluate the adjusted odds ratios (AOR) of factors associated with any tobacco use among men. The analysis included variables with p<0.005. Results are presented as AOR with 95% confidence intervals (CI) (Table 3). Age was a strong predictor, with older men having significantly higher odds of tobacco use. Analysis showed progressively higher odds with increasing age: men aged 19-24 (AOR: 2.29, 95% CI: 1.54–3.39), 25-29 (AOR: 2.69, 95% CI: 1.78–4.08), 30-39 (AOR: 3.09, 95% CI: 2.05–4.67), and 40-49 years (AOR: 4.28, 95% CI: 2.55–7.18). Marital status also played a role, with men who were widowed, divorced, or separated exhibiting higher odds of tobacco use (AOR: 2.52, 95% CI: 1.44–4.42), as compared to non-married men. Education was inversely associated with tobacco use; men with higher educational attainment, including complete secondary education (AOR: 0.25, 95% CI: 0.17–0.39) and higher education (AOR: 0.06, 95% CI: 0.03–0.13), had lower odds of tobacco use. Occupational factors also influenced tobacco use, with men in agricultural/domestic roles (AOR: 2.46, 95% CI: 1.49–4.06) and manual labor jobs (AOR: 2.62, 95% CI: 1.63–4.20) having significantly higher odds of tobacco use compared to those not working. Wealth was inversely associated with tobacco use, with men in the wealthiest quintile having 53% lower odds of tobacco use (AOR: 0.47, 95% CI: 0.33–0.68) compared to the poorest quintile. Mobile phone ownership, especially smartphones (AOR: 0.63, 95% CI: 0.48–0.81), was associated with lower odds of tobacco use. Geographically, men in coastal regions had 2.20 times higher odds of tobacco use compared to those in Phnom Penh (AOR: 2.20, 95% CI: 1.27–3.80). Alcohol consumption, were strongly associated with tobacco use; men who drank alcohol 1-5 days a month (AOR: 1.47, 95% CI: 1.20–1.80) or daily/almost daily (AOR: 3.88, 95% CI: 2.90–5.21) had significantly higher odds of tobacco use (see **Table 3**).

**Table 3.**
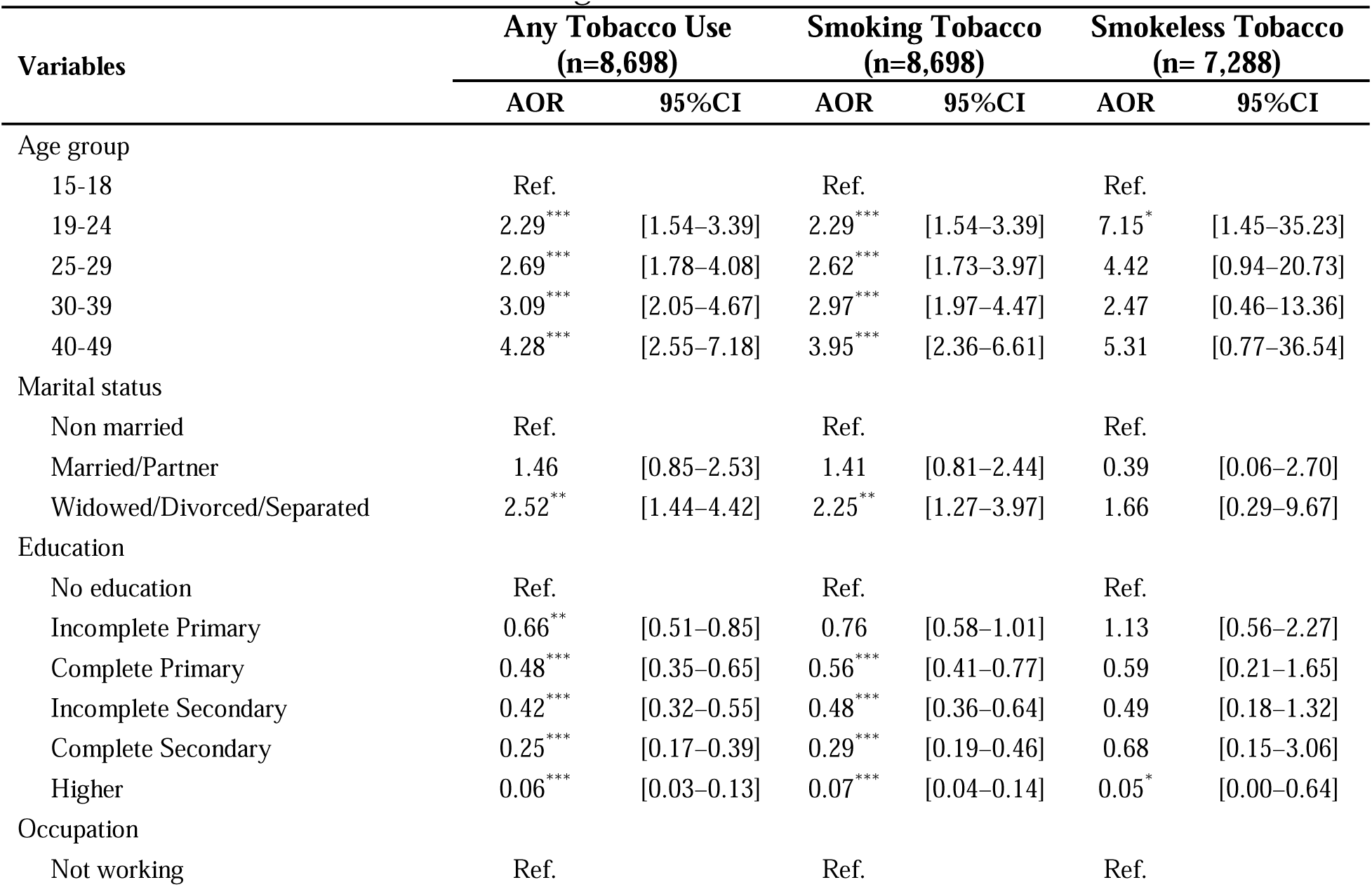
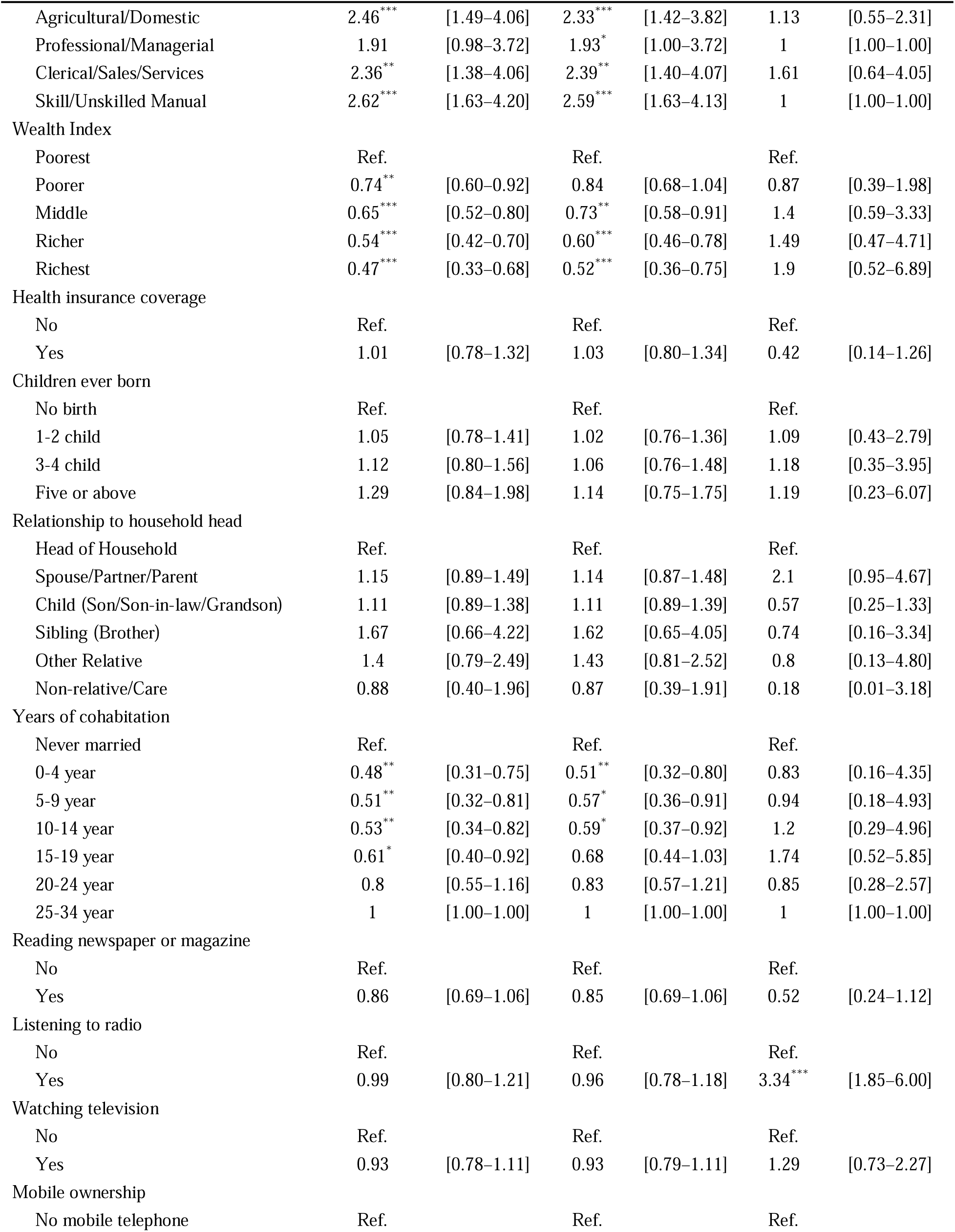
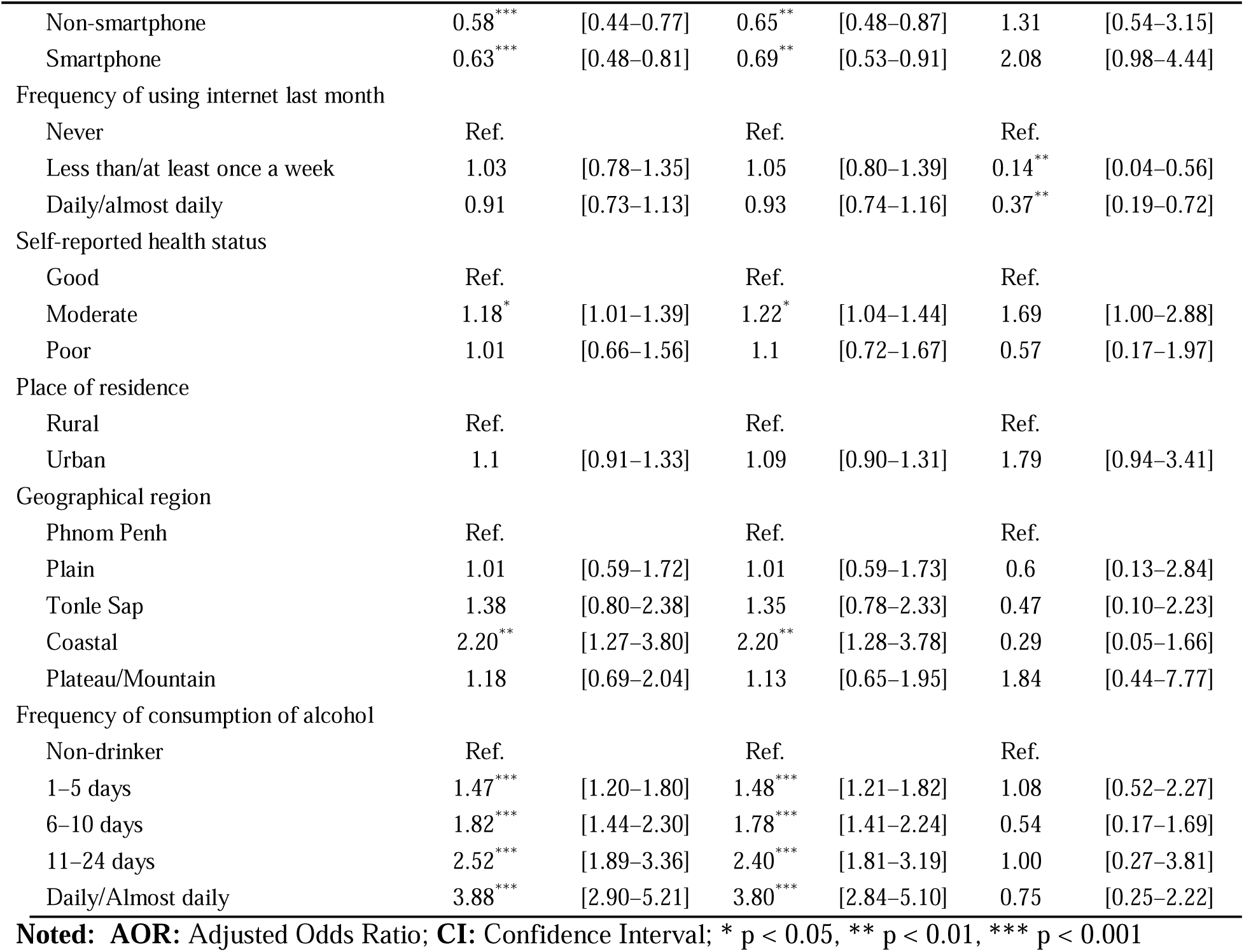
Adjusted Odds Ratios Evaluating Factors Associated with Any Tobacco Use, Smoking Tobacco, and Smokeless Tobacco among men.

| Variables | Any Tobacco Use<br>(n=8,698) |  | Smoking Tobacco<br>(n=8,698) |  | Smokeless Tobacco<br>(n= 7,288) |  |
| --- | --- | --- | --- | --- | --- | --- |
|  | AOR | 95%CI | AOR | 95%CI | AOR | 95%CI |
| Age group |  |  |  |  |  |  |
| 15-18 | Ref. |  | Ref. |  | Ref. |  |
| 19-24 | 2.29*** | [1.54–3.39] | 2.29*** | [1.54–3.39] | 7.15* | [1.45–35.23] |
| 25-29 | 2.69*** | [1.78–4.08] | 2.62*** | [1.73–3.97] | 4.42 | [0.94–20.73] |
| 30-39 | 3.09*** | [2.05–4.67] | 2.97*** | [1.97–4.47] | 2.47 | [0.46–13.36] |
| 40-49 | 4.28*** | [2.55–7.18] | 3.95*** | [2.36–6.61] | 5.31 | [0.77–36.54] |
| Marital status |  |  |  |  |  |  |
| Non married | Ref. |  | Ref. |  | Ref. |  |
| Married/Partner | 1.46 | [0.85–2.53] | 1.41 | [0.81–2.44] | 0.39 | [0.06–2.70] |
| Widowed/Divorced/Separated | 2.52** | [1.44–4.42] | 2.25** | [1.27–3.97] | 1.66 | [0.29–9.67] |
| Education |  |  |  |  |  |  |
| No education | Ref. |  | Ref. |  | Ref. |  |
| Incomplete Primary | 0.66** | [0.51–0.85] | 0.76 | [0.58–1.01] | 1.13 | [0.56–2.27] |
| Complete Primary | 0.48*** | [0.35–0.65] | 0.56*** | [0.41–0.77] | 0.59 | [0.21–1.65] |
| Incomplete Secondary | 0.42*** | [0.32–0.55] | 0.48*** | [0.36–0.64] | 0.49 | [0.18–1.32] |
| Complete Secondary | 0.25*** | [0.17–0.39] | 0.29*** | [0.19–0.46] | 0.68 | [0.15–3.06] |
| Higher | 0.06*** | [0.03–0.13] | 0.07*** | [0.04–0.14] | 0.05* | [0.00–0.64] |
| Occupation |  |  |  |  |  |  |
| Not working | Ref. |  | Ref. |  | Ref. |  |
|  | AOR | 95%CI | AOR | 95%CI | AOR | 95%CI |
| Agricultural/Domestic | 2.46*** | [1.49–4.06] | 2.33*** | [1.42–3.82] | 1.13 | [0.55–2.31] |
| Professional/Managerial | 1.91 | [0.98–3.72] | 1.93* | [1.00–3.72] | 1 | [1.00–1.00] |
| Clerical/Sales/Services | 2.36** | [1.38–4.06] | 2.39** | [1.40–4.07] | 1.61 | [0.64–4.05] |
| Skill/Unskilled Manual | 2.62*** | [1.63–4.20] | 2.59*** | [1.63–4.13] | 1 | [1.00–1.00] |
| Wealth Index |  |  |  |  |  |  |
| Poorest | Ref. |  | Ref. |  | Ref. |  |
| Poorer | 0.74** | [0.60–0.92] | 0.84 | [0.68–1.04] | 0.87 | [0.39–1.98] |
| Middle | 0.65*** | [0.52–0.80] | 0.73** | [0.58–0.91] | 1.4 | [0.59–3.33] |
| Richer | 0.54*** | [0.42–0.70] | 0.60*** | [0.46–0.78] | 1.49 | [0.47–4.71] |
| Richest | 0.47*** | [0.33–0.68] | 0.52*** | [0.36–0.75] | 1.9 | [0.52–6.89] |
| Health insurance coverage |  |  |  |  |  |  |
| No | Ref. |  | Ref. |  | Ref. |  |
| Yes | 1.01 | [0.78–1.32] | 1.03 | [0.80–1.34] | 0.42 | [0.14–1.26] |
| Children ever born |  |  |  |  |  |  |
| No birth | Ref. |  | Ref. |  | Ref. |  |
| 1-2 child | 1.05 | [0.78–1.41] | 1.02 | [0.76–1.36] | 1.09 | [0.43–2.79] |
| 3-4 child | 1.12 | [0.80–1.56] | 1.06 | [0.76–1.48] | 1.18 | [0.35–3.95] |
| Five or above | 1.29 | [0.84–1.98] | 1.14 | [0.75–1.75] | 1.19 | [0.23–6.07] |
| Relationship to household head |  |  |  |  |  |  |
| Head of Household | Ref. |  | Ref. |  | Ref. |  |
| Spouse/Partner/Parent | 1.15 | [0.89–1.49] | 1.14 | [0.87–1.48] | 2.1 | [0.95–4.67] |
| Child (Son/Son-in-law/Grandson) | 1.11 | [0.89–1.38] | 1.11 | [0.89–1.39] | 0.57 | [0.25–1.33] |
| Sibling (Brother) | 1.67 | [0.66–4.22] | 1.62 | [0.65–4.05] | 0.74 | [0.16–3.34] |
| Other Relative | 1.4 | [0.79–2.49] | 1.43 | [0.81–2.52] | 0.8 | [0.13–4.80] |
| Non-relative/Care | 0.88 | [0.40–1.96] | 0.87 | [0.39–1.91] | 0.18 | [0.01–3.18] |
| Years of cohabitation |  |  |  |  |  |  |
| Never married | Ref. |  | Ref. |  | Ref. |  |
| 0-4 year | 0.48** | [0.31–0.75] | 0.51** | [0.32–0.80] | 0.83 | [0.16–4.35] |
| 5-9 year | 0.51** | [0.32–0.81] | 0.57* | [0.36–0.91] | 0.94 | [0.18–4.93] |
| 10-14 year | 0.53** | [0.34–0.82] | 0.59* | [0.37–0.92] | 1.2 | [0.29–4.96] |
| 15-19 year | 0.61* | [0.40–0.92] | 0.68 | [0.44–1.03] | 1.74 | [0.52–5.85] |
| 20-24 year | 0.8 | [0.55–1.16] | 0.83 | [0.57–1.21] | 0.85 | [0.28–2.57] |
| 25-34 year | 1 | [1.00–1.00] | 1 | [1.00–1.00] | 1 | [1.00–1.00] |
| Reading newspaper or magazine |  |  |  |  |  |  |
| No | Ref. |  | Ref. |  | Ref. |  |
| Yes | 0.86 | [0.69–1.06] | 0.85 | [0.69–1.06] | 0.52 | [0.24–1.12] |
| Listening to radio |  |  |  |  |  |  |
| No | Ref. |  | Ref. |  | Ref. |  |
| Yes | 0.99 | [0.80–1.21] | 0.96 | [0.78–1.18] | 3.34*** | [1.85–6.00] |
| Watching television |  |  |  |  |  |  |
| No | Ref. |  | Ref. |  | Ref. |  |
| Yes | 0.93 | [0.78–1.11] | 0.93 | [0.79–1.11] | 1.29 | [0.73–2.27] |
| Mobile ownership |  |  |  |  |  |  |
| No mobile telephone | Ref. |  | Ref. |  | Ref. |  |
|  | AOR | 95%CI | AOR | 95%CI | AOR | 95%CI |
| Non-smartphone | 0.58*** | [0.44–0.77] | 0.65** | [0.48–0.87] | 1.31 | [0.54–3.15] |
| Smartphone | 0.63*** | [0.48–0.81] | 0.69** | [0.53–0.91] | 2.08 | [0.98–4.44] |
| Frequency of using internet last month |  |  |  |  |  |  |
| Never | Ref. |  | Ref. |  | Ref. |  |
| Less than/at least once a week | 1.03 | [0.78–1.35] | 1.05 | [0.80–1.39] | 0.14** | [0.04–0.56] |
| Daily/almost daily | 0.91 | [0.73–1.13] | 0.93 | [0.74–1.16] | 0.37** | [0.19–0.72] |
| Self-reported health status |  |  |  |  |  |  |
| Good | Ref. |  | Ref. |  | Ref. |  |
| Moderate | 1.18* | [1.01–1.39] | 1.22* | [1.04–1.44] | 1.69 | [1.00–2.88] |
| Poor | 1.01 | [0.66–1.56] | 1.1 | [0.72–1.67] | 0.57 | [0.17–1.97] |
| Place of residence |  |  |  |  |  |  |
| Rural | Ref. |  | Ref. |  | Ref. |  |
| Urban | 1.1 | [0.91–1.33] | 1.09 | [0.90–1.31] | 1.79 | [0.94–3.41] |
| Geographical region |  |  |  |  |  |  |
| Phnom Penh | Ref. |  | Ref. |  | Ref. |  |
| Plain | 1.01 | [0.59–1.72] | 1.01 | [0.59–1.73] | 0.6 | [0.13–2.84] |
| Tonle Sap | 1.38 | [0.80–2.38] | 1.35 | [0.78–2.33] | 0.47 | [0.10–2.23] |
| Coastal | 2.20** | [1.27–3.80] | 2.20** | [1.28–3.78] | 0.29 | [0.05–1.66] |
| Plateau/Mountain | 1.18 | [0.69–2.04] | 1.13 | [0.65–1.95] | 1.84 | [0.44–7.77] |
| Frequency of consumption of alcohol |  |  |  |  |  |  |
| Non-drinker | Ref. |  | Ref. |  | Ref. |  |
| 1–5 days | 1.47*** | [1.20–1.80] | 1.48*** | [1.21–1.82] | 1.08 | [0.52–2.27] |
| 6–10 days | 1.82*** | [1.44–2.30] | 1.78*** | [1.41–2.24] | 0.54 | [0.17–1.69] |
| 11–24 days | 2.52*** | [1.89–3.36] | 2.40*** | [1.81–3.19] | 1.00 | [0.27–3.81] |
| Daily/Almost daily | 3.88*** | [2.90–5.21] | 3.80*** | [2.84–5.10] | 0.75 | [0.25–2.22] |

**Noted:** AOR: Adjusted Odds Ratio; CI: Confidence Interval; \* $p < 0.05$ , \*\* $p < 0.01$ , \*\*\* $p < 0.001$

### Adjusted Odds Ratios Evaluating Factors Associated with Smoked Tobacco use

Odds of smoked tobacco use was higher in men aged 40–49 years (AOR: 3.95, 95% CI: 2.36– 6.61), 25–29 years (AOR: 2.62, 95% CI: 1.73–3.97), 30–39 years (AOR: 2.97, 95% CI: 1.97–4.47), and 19–24 years (AOR: 2.29, 95% CI: 1.54–3.39). Ever-married men had more than twice the odds of smoking (AOR: 2.25). Regarding education, men who received any level had lower odds with a decreasing trend as the number of years of education increased, as from primary (AOR: 0.56, 95% CI: 0.41–0.77), incomplete secondary (AOR: 0.48, 95% CI: 0.36–0.64), complete secondary (AOR: 0.29, 95% CI: 0.19–0.46), or higher education (AOR: 0.07, 95% CI: 0.04–0.14). Men have higher odds of smoking if they work in the agricultural/domestic (AOR: 2.33), professional/managerial (AOR: 1.93), clerical/sales/services (AOR: 2.39), and skilled/unskilled manual labor (AOR: 2.59). Men belonging to the middle (AOR: 0.73, 95% CI: 0.58–0.91), richer (AOR: 0.60, 95% CI: 0.46–0.78), and richest (AOR: 0.52, 95% CI: 0.36–0.75) were less likely to smoke. Alcohol consumption was strongly associated with smoking, with increasing trend as related to the number of days that men drank alcohol per month; on 1–5 days per month (AOR: 1.48, 95% CI: 1.21–1.82), on 6–10 days (AOR: 1.78, 95% CI: 1.41–2.24), on11–24 days (AOR: 2.40, 95% CI: 1.81–3.19), and daily/almost daily (AOR: 3.80, 95% CI: 2.84–5.10). Using a mobile phone (AOR: 0.65) or smartphones (AOR: 0.69, 95% CI: 0.53–0.91 was associated with reduced odds of smoking). Living in coastal regions had higher odds of smoking (AOR: 2.20, 95% CI: 1.28–3.78) (see **Table 3**).

### Adjusted Odds Ratios Evaluating Factors Associated with smokeless Tobacco

In contrast to smoked tobacco use, men aged 19-24 years were significantly more likely to use smokeless tobacco (AOR: 7.15, 95% CI: 1.45–35.23). However, similar to smoked tobacco use, higher education was significantly associated with reduced odds of smokeless tobacco use (AOR: 0.05, 95% CI: 0.00–0.64). Listening to the radio was strongly associated with increased smokeless tobacco use (AOR: 3.34, 95% CI: 1.85–6.00, p<0.001), while using the internet at least once a week (AOR: 0.14, 95% CI: 0.04–0.56, p<0.01) and daily/almost daily (AOR: 0.37, 95% CI: 0.19–0.72, p<0.01) were significantly associated with reduced smokeless tobacco use. Moderate self-reported health status was associated with higher odds of smokeless tobacco use (AOR: 1.69, 95% CI: 1.00–2.88), though this was borderline significant (see **Table 3**).

### Model Diagnostic for Multiple Logistic Regression Analysis

The goodness-of-fit statistics for the logistic regression models are presented in **S2 Table**. The models for any tobacco use (F(9, 652) = 1.40, p = 0.1861) and smoker status (F(9, 652) = 1.45, p = 0.1641) demonstrated acceptable fit. In contrast, the model for smokeless tobacco use showed poor fit (F(9, 652) = 22.42, p < 0.001), suggesting that important predictors may be missing or that alternative model specifications are needed (**S2 Table**).

To assess the models’ discriminatory power, Receiver Operating Characteristic (ROC) curves were generated. The area under the curve (AUC) values indicated good discrimination for the any tobacco use and smoker status models, while the smokeless tobacco use model had a lower AUC, consistent with the poor model fit (**Fig 2**).

**Fig 2.**
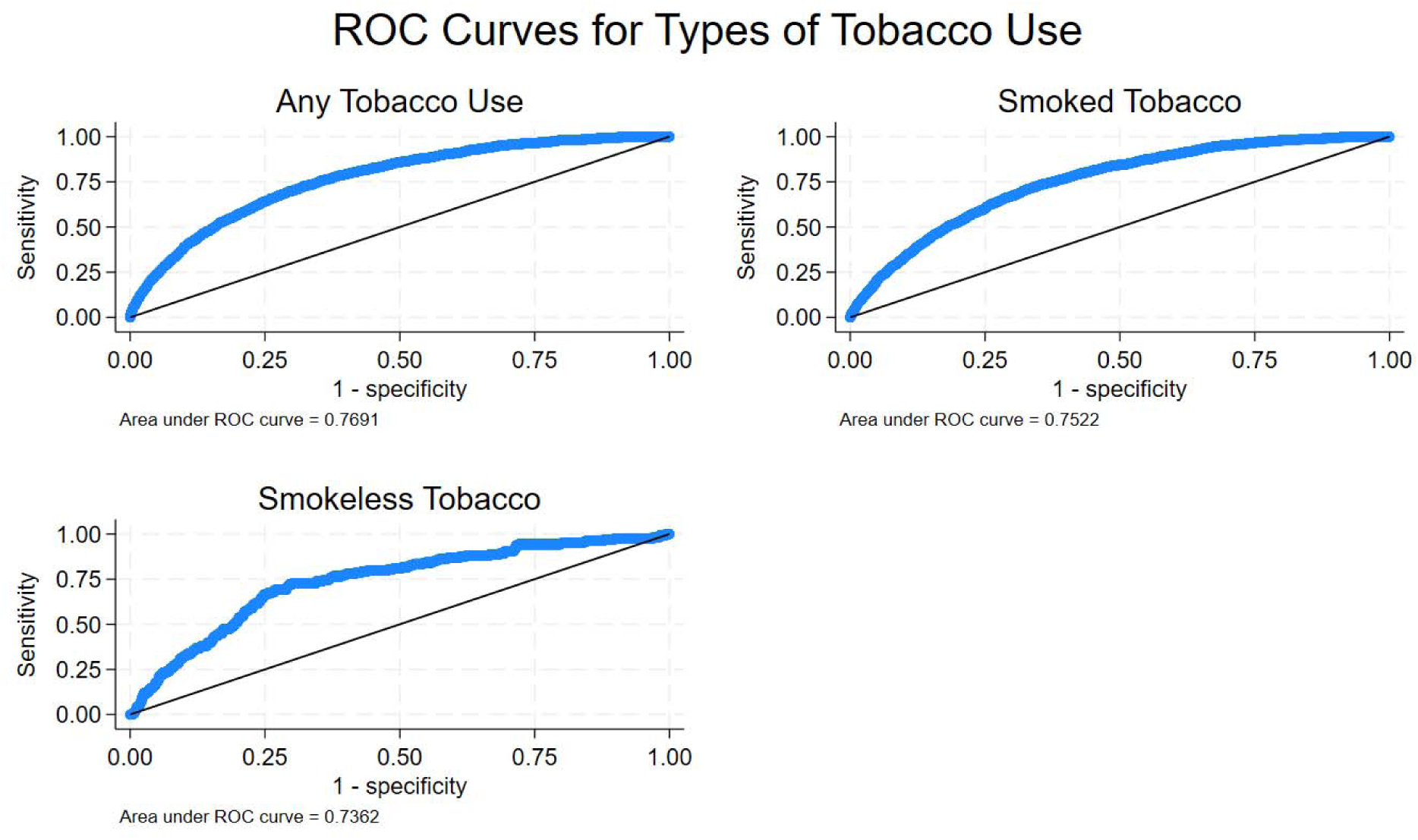
Receiver Operating Characteristic (ROC) curves for logistic regression models of tobacco use types.

### Interaction analyses on the factors associated with Any Tobacco Use

Several interaction effects were identified in relation to tobacco use among men aged 15–49 years.

A significant interaction was found between age and education. The predicted probability of tobacco use increased with age, particularly among men with no formal education or only primary education. In contrast, those with secondary or higher education showed consistently lower probabilities of tobacco use across age groups (**Fig. 3A**). An interaction between age and alcohol consumption was also evident. Among men who consumed alcohol, older age groups had a markedly higher predicted probability of tobacco use compared to younger men. This suggests that the combined influence of alcohol use and increasing age contributes to greater tobacco use risk (**Fig. 3B**). The association between education and occupation further revealed that men with lower education working in manual or agricultural jobs were more likely to use tobacco compared to those in professional or service occupations with higher education (**Fig. 3C**). Lastly, mobile phone ownership and internet use showed a notable interaction. Men who owned a mobile phone but rarely or never used the internet had higher predicted probabilities of tobacco use than those who used the internet regularly, highlighting possible disparities in exposure to health information and behavior change messaging (**Fig 3).**

**Fig 3.**
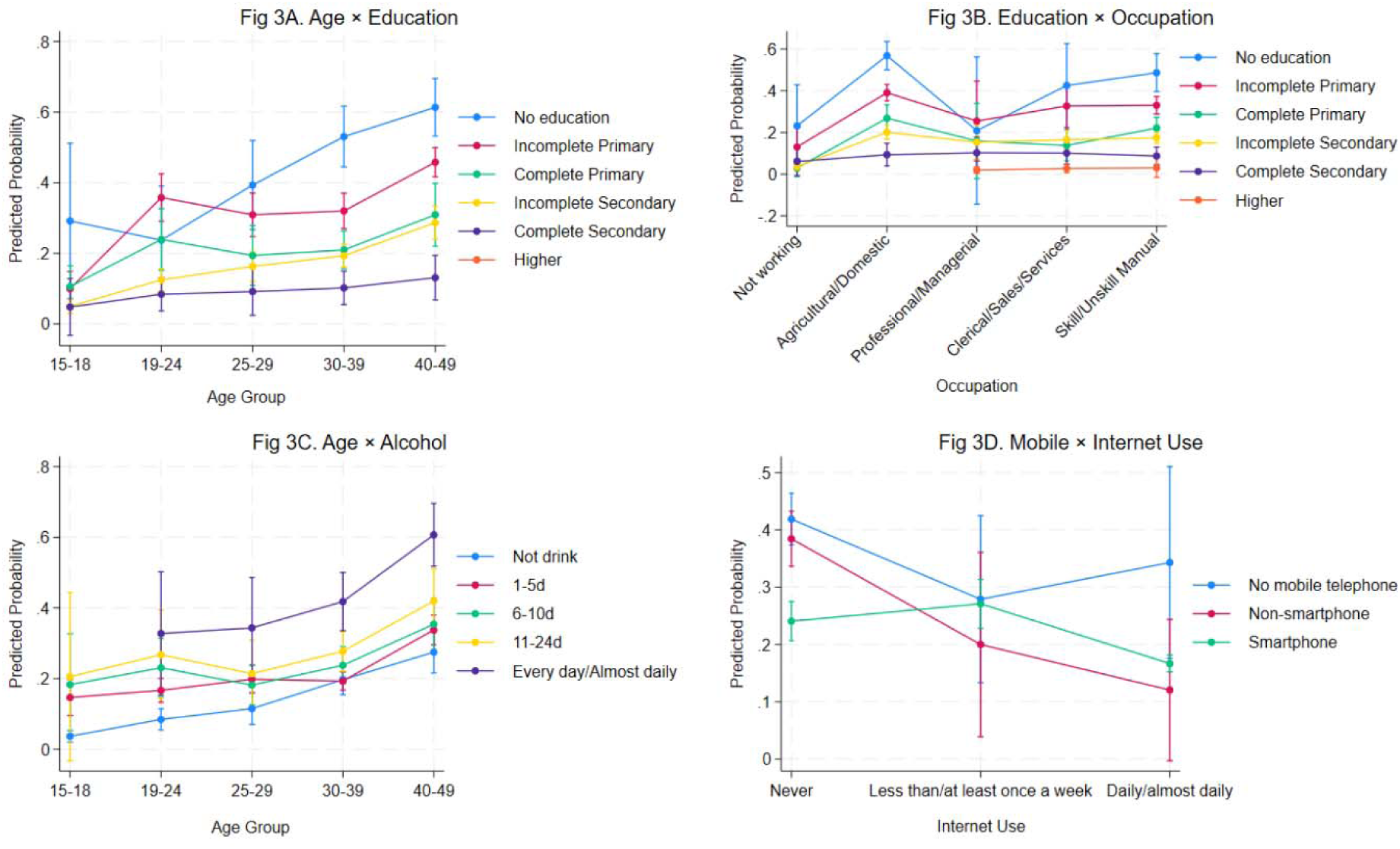
Predicted probabilities of any tobacco use by interaction effects: (Fig. 3A) Age and Education; (Fig. 3B) Age and Alcohol Consumption; (Fig. 3C) Education and Occupation; (Fig. 3D) Mobile Phone Ownership and Internet Use.

## Discussion

This study aimed to identify socio-demographic and behavioral factors, including alcohol use, associated with any tobacco use, smoked tobacco use, and smokeless tobacco use among men 15-49 years in Cambodia. The overall prevalence of tobacco use in any form was found to be 21.5%, with 20.1% of men reporting smoking tobacco products, and 0.93% using smokeless tobacco. There was a notable geographical variation in tobacco use, with the lowest prevalence observed in Phnom Penh and the highest in mountainous regions. These highlight the continued significance of tobacco consumption as a major public health issue in Cambodia, particularly in rural and remote areas where tobacco use may be more prevalent. Several significant predictors of any tobacco use, smoking tobacco and smokeless tobacco, including age, education, occupation, and wealth, highlight the multifactorial nature influencing tobacco use in this population. Most of the predictors are significant social determinants of health. Unsurprisingly, alcohol consumption is strongly associated with tobacco use across all forms. The strong association between alcohol consumption and tobacco use is a critical finding, as co-use of alcohol and tobacco increases the overall health risks for individuals, particularly for chronic diseases, including diabetes, hypertension, lung cancers and chronic respiratory diseases, cardiovascular disorders, liver disease, and accidental traffic injuries [20,30,31]. Integrated tobacco and alcohol control policies are essential in Cambodia [32–35]. Addressing both risk factors simultaneously in public health campaigns, cessation programs, and regulatory measures can be an effective and efficient approach in reducing overall substance use. Additionally, healthcare providers should be trained to recognize the co-occurrence of these behaviors and provide integrated support for cessation [6].

Our findings are slightly lower than the Global Tobacco Control Report for Cambodia 2023, where 28.6% of men aged 15 and above use tobacco, of whom 28.4% smoke tobacco, and 0.3% use smokeless tobacco [36]. According to the Cambodia NATS, in 2021, cigarette smoking prevalence relatively decreased among men from 32.3% in 2014 to 25.37% in 2021 [17]. However, the Cambodia STEPS survey 2023 was higher with 35.5% of men aged 18-69 years reporting current use of any tobacco, with 29.4% smoking tobacco products, and 0.7% using smokeless tobacco [16]. Among the STEPS survey respondents, 12.3% of men aged 18-69 had quit smoking, of whom one in four men aged 45-59 and one in three aged 60-69 reported being previous daily smokers [16]. Importantly, Cambodia has not yet achieved its national target between 2018-2027 of a 30% relative reduction in the prevalence of current tobacco use among adults aged 15 and above [16]. In Cambodia, factors such as limited health awareness, lower education levels, and less effective tobacco control measures contribute to higher tobacco use. This is consistent with trends observed in other South and Southeast Asian countries [37–39]. On the other hand, smokeless tobacco use was generally low across most provinces, with the highest rates seen in Mondulkiri (19.1%) and Stung Treng (6%). This aligns with regional studies that indicate smokeless tobacco use is prevalent among specific ethnic communities [38,40].

While age is the only non-modifiable risk factor for tobacco use, most of the identified predictors—such as education, occupation, and household wealth index—are significant social determinants of health [7,8,16,20]. Lower education levels and engagement in manual labor or agricultural occupations were linked to a higher prevalence of tobacco use. Men from poorer households were more likely to use tobacco compared to those from wealthier backgrounds, reinforcing the notion that socioeconomic inequalities play a crucial role in health risk behaviors [1,8]. These findings suggest that interventions should focus on reducing socio-economic disparities by promoting education and economic empowerment as key strategies in tobacco as well as alcohol control.

This study highlights the significant association between tobacco use and alcohol consumption among Cambodian men aged 15–49 years. The findings reinforce well-documented evidence that tobacco and alcohol use frequently co-occur, with alcohol consumption emerging as one of the strongest predictors of tobacco use [21,41–43]. The odds of tobacco use increase with the frequency of alcohol consumption, indicating a dose-response relationship. This association underscores the need for integrated policy interventions targeting both risk factors simultaneously, including integrated approaches to behavioral risk factor reduction. Although Cambodia has made progress in tobacco control through the implementation of the Tobacco Control Law and the ratification of the WHO Framework Convention on Tobacco Control (FCTC), policies addressing alcohol consumption remain less developed [15,34,44]. Policy-makers can take inspiration from the more successful strategies recommended in the FCTC for alcohol control. Furthermore, given that alcohol consumption increases the likelihood of tobacco use, future public health policies integrating both risk factors in prevention and cessation programs can prove to be more efficient [45].

Comprehensive behavioral interventions should be developed to address both tobacco and alcohol use concurrently. Smoking cessation programs can incorporate components targeting alcohol reduction, while alcohol harm reduction initiatives can integrate tobacco cessation support [35,46]. Stricter regulations should be placed on the marketing and advertisement of both tobacco and alcohol, particularly in media that targets youth and low-income populations. Lessons can be drawn from tobacco control policies, such as restrictions on advertising, plain packaging, and taxation strategies, and adapted for alcohol control [33,34,44]. Increasing excise taxes on both tobacco and alcohol products can serve as a deterrent to consumption, particularly among low-income groups. Higher taxation has been an effective strategy in reducing tobacco use and could similarly reduce alcohol consumption [34,44].

## Policy Implementation

The Royal Government of Cambodia has introduced several laws and policies in order to control tobacco. In 2006, the Tobacco Product (Control and Regulation) Act 2006 was introduced, which prohibited smoking in public places and set limits to the advertisement and promotion of tobacco [34,44]. Strengthening enforcement of Cambodia’s tobacco control laws, including taxation, advertising restrictions, and smoke-free policies, is essential to curbing use, especially in rural areas [46]. Given the strong association between low education and high tobacco use, integrating anti-tobacco education into school curricula and community health programs could help mitigate the problem [44]. Furthermore, targeted tobacco control programs, such as mobile health (mHealth) interventions, may also be beneficial [14]. Special attention should be given to addressing the intersection of tobacco and alcohol use. By leveraging on the existing tobacco control framework and extending similar policies to alcohol regulation, Cambodia can advance its public health goals, reduce the burden of non-communicable diseases, and improve overall health outcomes.

## Strengths and limitations

The current study has a number of strengths. This is based on a population-based household survey as a national-level study, which has a validated questionnaire and a high response rate (97%) of men aged 15–49 years in Cambodia to estimate the prevalence of tobacco status. As such, the findings of the study are generalizable to the target population. Moreover, the probability of the existence of measurement error in the study is lower compared to any other cross-sectional survey of the Cambodian population because of the utilization of standard and validated measurement tools by the DHS program. However, the study has some limitations. It only examines data from male respondents; hence, tobacco use among women was not analyzed. The temporal relationship between the social determinants, alcohol consumption, and the tobacco variable cannot be established in a cross-sectional study. Furthermore, tobacco and alcohol consumption were collected as self-reported without asking explicitly about the consumed amount; hence, there is a chance of being subject to reporting bias, potentially underestimating due to social desirability.

## Conclusion

The current study has shown that 21% of Cambodian men currently tobacco, 20.1% smoke tobacco, and 0.93% smokeless tobacco. Men having lower education, lower socioeconomic status, from the remote provinces, and with increasing age were more likely to consume tobacco and also have high alcohol consumption [21]. There is an urgent need to implement the recently endorsed Tobacco Control and Regulation Act in Cambodia, which has highlighted major tobacco control activities, and to the extent possible, incorporate alcohol control. Efforts should be focused on creating perceptions of cultural unacceptability of tobacco use - either chewing or smoking – and alcohol use among the Cambodian population. Regulations regarding alcohol advertising, sales, and consumption have recently been enforced; implementation can be patterned after the Tobacco Control and Regulation Act.

## Acknowledgments

The authors would like to thank DHS-ICF, who approved the data used for this paper.

## Data Availability

This study used the 2021-2022 Cambodia Demographic and Health Survey (CDHS) datasets. The DHS data are publicly available from the website at (URL:https://www.dhsprogram.com/data/available-datasets.cfm).

## Abbreviations

ACS: American Cancer Society
AOR: adjusted odds ratio
CDHS: Cambodia Demographic Health Survey
EA: enumeration areas
FCTC: Framework Convention on Tobacco Control
LMICs: low- and middle-income countries
IHD: ischemic heart disease
NATS: National Adult Tobacco Survey
PPS: probability proportional to size
WHO: World Health Organization
SDGs: Sustainable Development Goals

---

**Table S1.**
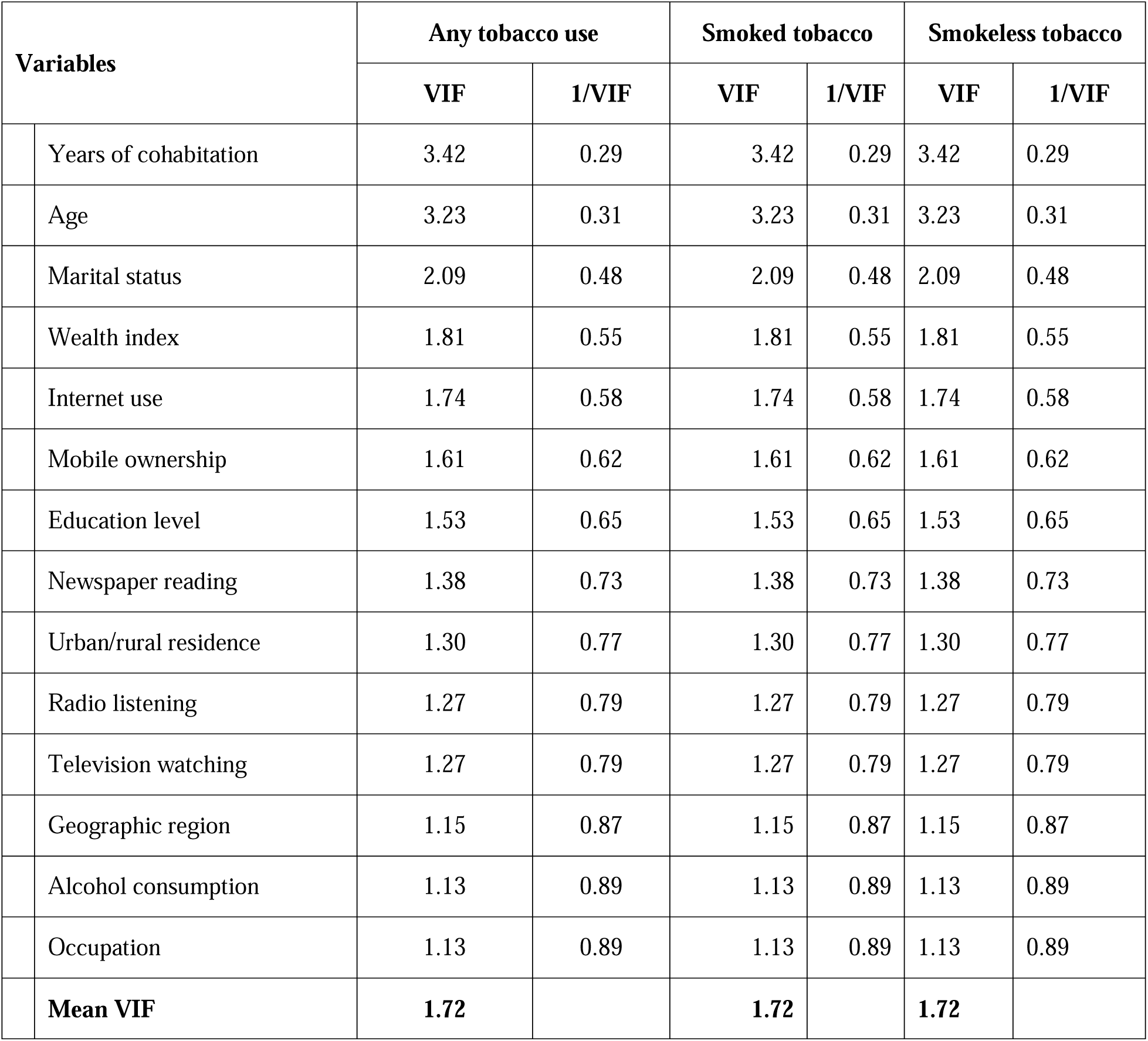
Variance Inflation Factors (VIFs) for Variables in Logistic Regression Models of Tobacco Use among Cambodian Men.

**Table S2.** Goodness-of-Fit Statistics for Multiple Logistic Regression Models.

| Outcome Variable | F Statistic (df1, df2) | p-value | Model Fit |
| --- | --- | --- | --- |
| Any Tobacco Use | F(9, 652) = 1.40 | 0.1861 | Acceptable Fit |
| Smoker Status | F(9, 652) = 1.45 | 0.1641 | Acceptable Fit |
| Smokeless Tobacco Use | F(9, 652) = 22.42 | 0.0000 | Poor Fit |

